# A Natural Language Processing Pipeline based on the Columbia-Suicide Severity Rating Scale

**DOI:** 10.1101/2024.12.19.24319352

**Authors:** Lauren A. Lepow, Prakash Adekkanattu, Marika Cusick, Hilary Coon, Brian Fennessy, Shane O’Connell, Charlotte Pierce, Jessica Rabbany, Mohit Sharma, Mark Olfson, Amanda Bakian, Yunyu Xiao, Niamh Mullins, Girish N. Nadkarni, Alexander W. Charney, Jyotishman Pathak, J. John Mann

## Abstract

**Importance:** Diagnostic codes in the Electronic Health Record (EHR) are known to be limited in reporting patient suicidality, and especially in differentiating the levels of suicide severity.

**Objective:** The authors developed and validated a portable natural language processing (NLP) algorithm for detection of suicidal ideation (SI) and suicide-related behavior and attempts (SB/SA) in EHR data. The algorithm was then deployed, and SI and SB/SA ascertainment was compared to that of International Statistical Classification of Diseases (ICD-9 and 10) diagnostic codes.

**Design:** A group of experts designed the pipeline to detect and distinguish suicide severity based on the Columbia-Suicide Severity Rating Scale (C-SSRS). Notes were manually annotated to create the “Gold Standard” with which the algorithm output was evaluated for accuracy.

**Setting:** The algorithm was developed at two academic medical centers, Weill Cornell Medicine (WCM), the Mount Sinai Health System (MSHS), and tested at these two, plus a third, the University of Utah Healthcare Center (UUHSC).

**Participants:** Notes were from participants with psychiatric encounters at the three institutions.

**Main Outcomes:** The two main outcomes were the accuracy scores of the NLP pipeline and comparison of ascertainment rates to ICD codes.

**Results:** F1 accuracy scores ranged from 0.86-0.97 at the three sites. The NLP rate of detection of SB/SA was almost 30 times higher, and SI was almost 10 times higher, when compared with that of diagnostic codes. NLP detected almost all cases detected by diagnostic codes. No bias in performance was found for race/ethnicity and performance was comparable in psychiatric and non-psychiatric EHRs.

**Conclusions and Relevance:** EHRs from cohorts with psychiatric diagnoses or encounters at WCM, MSHS, and UUHSC had SI and SB/SA extracted using an NLP algorithm based on parameters defined by the C-SSRS. Validity was determined by comparing the algorithm output to manual annotations of clinical notes by domain experts. NLP-detection of SI and SB/SA was compared with that of ICD codes across a range of demographic groups. Algorithm performance was also examined for bias in minoritized groups and in non-psychiatric notes.

**KEY POINTS:** *Question:* Can we automate the extraction of data available in clinical notes to accurately detect and distinguish patients with suicidal ideation (SI) and suicidal behavior (SB)?

*Findings:* Our Natural Language Processing (NLP) approach was able to identify and distinguish SI and SB at three different hospital systems with benchmarked accuracy scores (above 0.85). The rate of detecting SI and SB using the algorithm was 10-30 times that of diagnostic codes found in the Electronic Health Record.

*Meaning:* Our algorithm renders the use of International Classification of Disease (ICD) diagnostic codes for SI and SB ascertainment obsolete.

## Introduction

Suicide attempts (SAs), as defined in the Columbia Suicide Severity Rating scale, are the strongest known risk factors for future suicide attempts (1–3). To a lesser degrees, suicide-related behaviors and suicidal ideation also predict risk of future suicide attempts. The term *suicidality* encompasses all three phenomena but blurs the distinction that the rank order of prediction of future suicide attempts is SA>SB>SI (4). Therefore, distinguishing suicide attempts and suicide-related behaviors from ideation allows for more precision for prevention, prognostics, clinical decisions, and research strategies. The Columbia—Suicide Severity Rating Scale (C-SSRS) was designed to help the clinician systematically determine the presence and severity of SI, SB, and SA (4). In mental health settings, clinicians are often trained on the C-SSRS to assess and document patient suicide risk (4). Suicide prevention strategies outlined by the Centers for Disease Control and Prevention (CDC), the U.S. Department of Health and Human Services, and the National Institutes of Health are based on establishing the severity of past and current suicidality (5). However, prior EHR-based research suggested that structured diagnostic codes are inadequate for identifying SI and SB (4–6,10). For example, only 3% of 1025 patients with suicide ideation recorded in clinical notes had an International Classification of Disease (ICD)-9 code for SI (10). Utilizing existing free-text clinical notes from electronic health records (EHRs) for identifying patients with suicide risk provides additional data for detection of these higher risk patients. The fact that 80% of people who died by suicide had a physician visit in the year prior to death–and 45% in the previous month–underscores the importance of optimizing EHR data in the assessment of suicide risk (3, 4).

Natural language processing (NLP) approaches have previously been designed and tested for extracting suicidality information from EHRs; these methods range from rule-based algorithms using a dictionary of terms to machine learning methods (6–15). Most NLP algorithms were developed at single institutions, and therefore portability to other healthcare systems is generally unknown. Cross-site validation of two NLP methods developed independently at King’s College, London (KCL) and Weill Cornell Medicine (WCM), without direct input from the other site in algorithm development, failed to achieve comparable cross-institution performance (16). A second challenge is that studies find diagnostic biases in minoritized groups (11, 12) including in EHRs (10); therefore, it is important that performance metrics of NLP systems are evaluated for bias. The aim of the present study was to achieve comparable performance across the three participating sites with varied care settings, documentation practices, and patient populations.

Building on the WCM lexicon-based approach previously developed, this new algorithm was developed collaboratively at WCM and the Mount Sinai Health System (MSHS) and then tested at both sites. Additional and independent performance evaluation of the algorithm was conducted at a third site— the University of Utah Healthcare Center (UUHSC). This NLP was designed, using the C-SSRS, to identify the separate risk categories of suicide attempt/suicide-related behaviors and suicidal ideation, and to exclude non-suicidal self-harm. Validity of the NLP results was established by comparing the NLP findings to manual clinical note assessments by two raters blind to the NLP results and each other’s findings. Differences of opinion were adjudicated by domain experts and reviewed by JJM, one of the creators of the C-SSRS. Serviceability in other healthcare systems was demonstrated by testing the algorithm across three different sites. Performance across different demographic groups evaluated potential bias.

Finally, performance was evaluated in psychiatric versus non-psychiatric notes to explore generalizability. The validated algorithm was deployed in a much larger cohort of patients in two of the sites with more diverse patient populations to compare the NLP performance to diagnostic codes and across different demographics to detect any bias effect.

## Materials and Methods

### Data Sources

The NLP algorithm was developed at WCM and MSHS and validated at three academic medical centers, WCM, MSHS, and UUHSC. See Supplement 1 for more details of the three underlying cohorts from which the test-set of notes was sampled.

### Development of the NLP algorithm

With input from co-author JJM, a developer of the C-SSRS, lexicons were developed by our team of clinicians, epidemiologists, and data scientists. Lexicons were developed separately for SI and for SB/SA. The entire range of severity of SI, as defined by the C-SSRS, was considered: wish to be dead at the least severe end to suicidal ideation with both intent and plan as the most severe. SB/SA was based on the C-SSRS behavior subscale and included actual, aborted, and interrupted suicide attempts as well as preparatory behavior. This category was defined by the C-SSRS as a nonfatal self-directed behavior with any intent to die that may or may not result in injury. Different from other suicidality NLP pipelines, non-suicidal self-injurious behavior such as cutting or burning oneself without the intent to die, which is a distinct phenomenon, was excluded from SB (5, 12). Preparatory behavior was defined by the C-SSRS as anything beyond a verbalization or thought of suicide, such as assembling a method (e.g., collecting pills, acquiring a firearm or piece of rope, etc.) or preparing for one’s death by suicide (e.g., writing a suicide note, giving away valuable possessions, writing a will etc). See Figure 1 for a schematic of the NLP development.

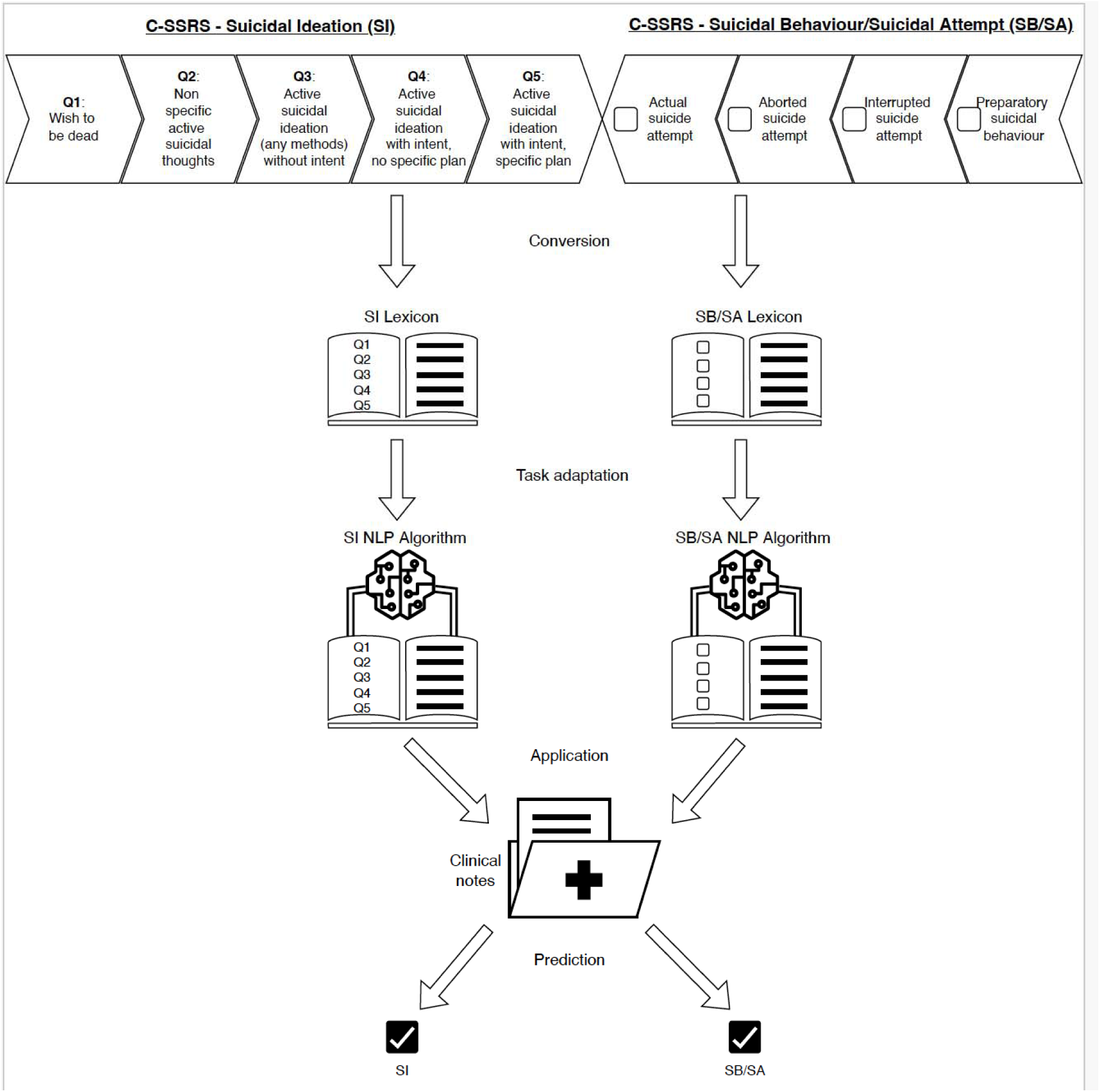
Schematic of study workflow. The Columbia Suicide Severity Rating Scale (C-SSRS) was used to design lexicons for suicidal ideation (SI, left in figure) and suicidal behavior/suicidal attempt (SB/SA, right in figure). SI and SB/SA lexicons, along with the NegEx algorithm (17), make up the rules-based natural language processing (NLP) algorithm to create two classification models: one for classifying SI, and another for classifying SB/SA.

The lexicon approach was paired with the NegEx (17) algorithm, which identifies negated and off-target findings in clinical text. NegEx uses additional lexicons, “modifiers,” that negate the suicidality term as an affirmative patient descriptor (e.g., “not”, “family history of”). The full set of modifiers is provided on our GitHub repository. An encounter note was considered “positive” for SI or SB/SA if at least one affirmative mention was documented during that clinical encounter. Annotation guidelines were created to match the lexicons. Notes were labeled with each target phrase tagged as SI, SI with plan, SI with intent, SI without plan, SI without intent, SA term, and SB term and options to denote the context of the term as negated, historic, and unrelated. For the purposes of this study, during the evaluation phase, SI with or without plan and intent were grouped and considered as one category; SA and SB were also grouped. Furthermore, past and current suicidality was not distinguished. At each site, inter-rater agreement was calculated using Cohen’s Kappa statistic (Supplemental Table S1). Because the underlying NLP pipeline had already been established (16), only limited modifications were required. For site-specific modifications to the lexicons, see Supplement 2.

### Validation by comparison with manual annotation

To create a corpus of notes for the manual annotation validation procedure at each site, instances of suicidality were enriched in the note selection process. Hence, about half the notes were selected from “cases,” or patients with ICD evidence of suicidality, and “controls,” who lacked ICD evidence of suicide symptoms. To increase the diversity of language style, the number of note-writers was maximized and the amount of copy-forwarded text was minimized by selecting a maximum of 20 notes from any one patient. The exact criteria for cases and controls varied at each institution (as detailed below) adding inter-site variability into the annotated set. At WCM, notes were filtered to include “psychiatry encounters” and then 200 notes were taken from patients with an ICD diagnosis of SB/SA (n=16) and 200 notes from patients without SA/SB (n=19). At MSHS, notes were filtered by the inclusion of the phrase “history of present illness” to select for clinician evaluations. The filtering resulted in 25 cases with these note types who also had an ICD code for any suicidality (SI and/or SB/SA) and 194 notes. 21 patients without SI and SB/SA ICD codes and 135 notes were randomly selected as controls. At UUHSC, 293 notes were chosen randomly from the 22 suicide deaths with ICD codes indicating prior nonlethal suicidality (“cases”), and 111 notes were randomly selected from the 22 age-/sex-/race-matched living individuals with no ICD codes for suicidality (“controls”).

Instances of suicidality were converted to a document-level classification such that each note was labeled as SI present (1) or absent (0) and SB/SA 1 or 0. At each site, NLP-predicted document-level classification was compared to the annotated note set. Each note was read by two raters independently and scored for SA/SB or SI. When the two raters disagreed a third party or senior author JJM adjudicated. F-scores are used as test accuracy summary statistics: F-1 equally considers precision and recall whereas F-2 places more emphasis on recall, therefore penalizing the score more heavily when the algorithm misses a mention that the manual annotators found. The macro-averages of the classification classes (0 and 1) are calculated for precision, recall, and F-scores for SI and SB/SA.

### Comparison of NLP and diagnostic codes in detection of SA/SB and SI in large EMR data sets from two different medical centers

At WCM, the algorithm was deployed in a cohort of 14,462 patients with at least one psychiatric encounter for a total of 147,631 notes. At MSHS, the algorithm was deployed in the acute care corpus described above, totaling 286,459 notes from 33,797 patients. The prevalence of ICD codes for suicidality was compared to that of the NLP algorithm. ICD codes for SI and SB/SA were reviewed from a list of 10,205 ICD-9 and ICD-10 codes to remove irrelevant codes like poisoning and self-harm. ICD and NLP ascertainment of SI and SB/SA were compared among different demographic and diagnostic groups. Patients were also divided into groups of low, medium, and high numbers of notes in the corpus to observe whether ICD-based diagnosis versus NLP-derived mentions of SI and SB/SA prevalence varied across these groups.

### Testing performance accuracy across race and ethnicity groups

For further validation, at two of the sites (WCM, MSHS) with notable diversity, the accuracy metrics listed above were stratified by race and ethnicity. Given the relatively small sample size of patients in the gold-standard corpus, race was grouped into white and non-white, and ethnicity into Hispanic/Latinx and non-Hispanic/Latinx.

### Testing performance accuracy outside psychiatric encounters

At UUHSC, notes were stratified into psychiatric and non-psychiatric encounters to see whether accuracy was maintained in non-psychiatric notes.

## Results

Demographics of the cohorts are described in Table 1.

**Table 1:** Demographic Characteristics of Patients that Comprise Manual Annotation.

| Site<br>(number of patients) |  | WCM<br>(n=32) | MSHS<br>(n=46) | UUHSC<br>(n=44) |
| --- | --- | --- | --- | --- |
| Age (years) | <18 | 1 | 12 | 0 |
|  | 18-39 | 17 | 16 | 23 |
|  | 40-59 | 7 | 15 | 21 |
|  | 60+ | 7 | 3 | 0 |
| Sex | Female | 22 | 26 | 19 |
|  | Male | 10 | 20 | 25 |
| Race and Ethnicity | White | 18 | 10 | 44 |
|  | Non-white | 14 | 36 | 0 |
|  | Hispanic/Latinx | 2 | 9 | 1 |
|  | Non-Hispanic/Latinx | 16 | 18 | 37 |
| ICD diagnosis of suicidality | Yes | 16 | 24 | 22 |
|  | No | 16 | 22 | NA |
| Suicide death | Yes | NA | NA | 22 |
|  | No | NA | NA | 22 |

*WCM refers to Weill Cornell Medicine, MSHS refers to Mount Sinai Health System, and UUHSC refers to University of Utah Healthcare Centre.*

### NLP algorithm validity

The inter-rater agreement among manual annotators was robust across all three sites, as detailed in Appendix Table 2. When evaluated against the manually annotated free text notes as the gold standard, macro-averaged (for class 0 and 1 as described in Methods) precision and recall statistics were produced for the NLP algorithm: At WCM, the algorithm correctly identified 45 out of 63 notes that mentioned SI for a precision of 0.85 and a sensitivity of 0.93; it correctly identified 115 out of 125 notes that mentioned SB/SA for a precision of 0.91 and sensitivity of 0.83. At MSHS, the algorithm correctly identified 196 out of 229 notes that mentioned SI for a precision of 0.85 and a sensitivity of 0.89. For SB/SA, it correctly identified 139 out of 161 notes for both a precision and sensitivity of 0.86. At UUHSC, the algorithm correctly identified 200 of 211 notes with SI with a low false negative rate of only 6 notes, for a precision of 0.95 and a sensitivity of 0.97. For SB/SA, the algorithm correctly identified 138 out of 151 notes again with a low false negative rate of only 1 note, resulting in a precision of 0.95 and sensitivity of 0.99. The macro-average F1-scores achieved by the NLP algorithm ranged from 0.92 to 0.96 for SI and from 0.86 to 0.97 for SB/SA (Table 2).

**Table 2:**
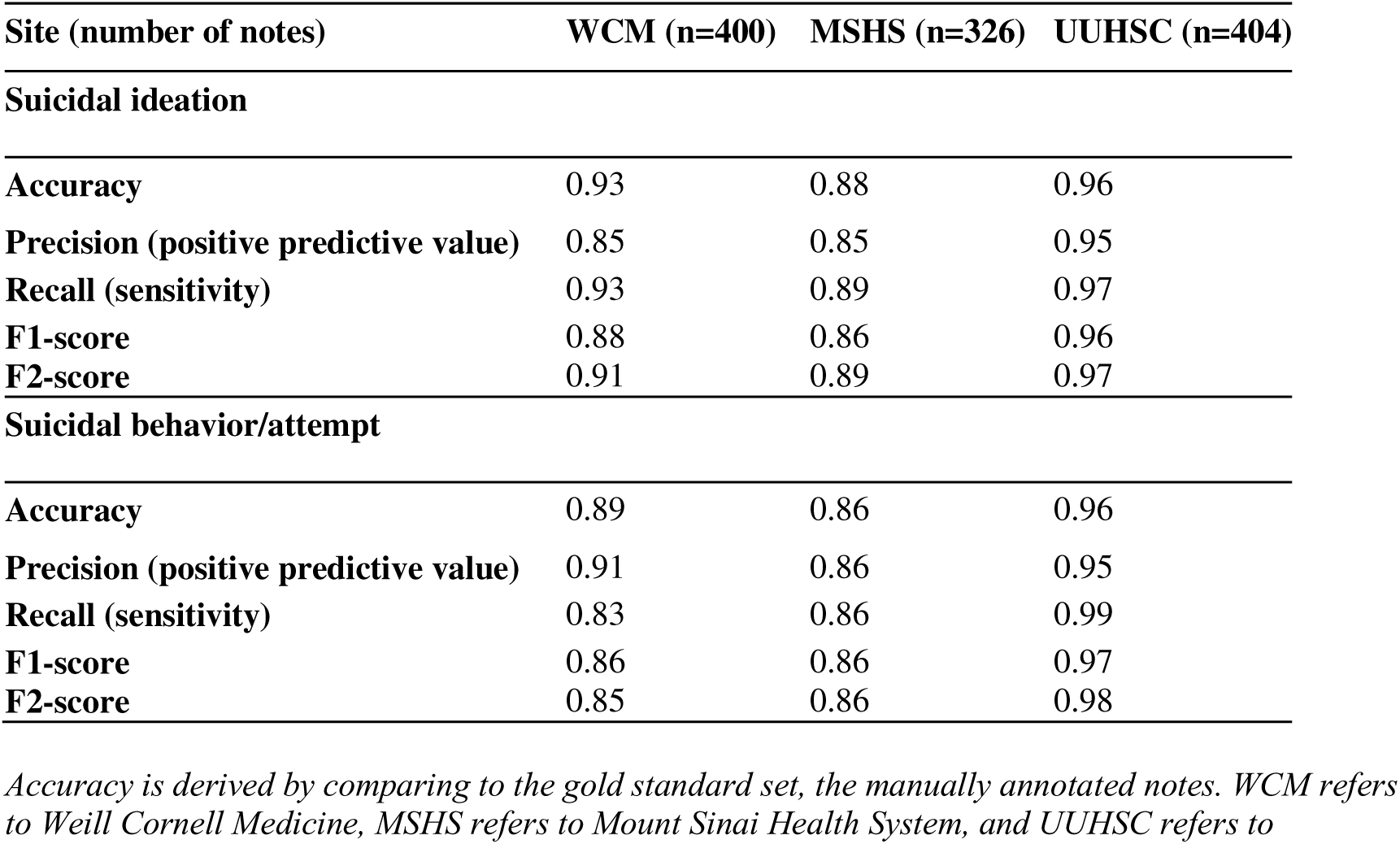
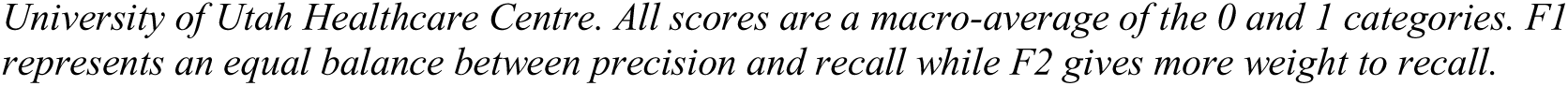
Accuracy metrics of the NLP algorithm.

| Site (number of notes) | WCM (n=400) | MSHS (n=326) | UUHSC (n=404) |
| --- | --- | --- | --- |
| **Suicidal ideation** |  |  |  |
| Accuracy | 0.93 | 0.88 | 0.96 |
| Precision (positive predictive value) | 0.85 | 0.85 | 0.95 |
| Recall (sensitivity) | 0.93 | 0.89 | 0.97 |
| F1-score | 0.88 | 0.86 | 0.96 |
| F2-score | 0.91 | 0.89 | 0.97 |
| **Suicidal behavior/attempt** |  |  |  |
| Accuracy | 0.89 | 0.86 | 0.96 |
| Precision (positive predictive value) | 0.91 | 0.86 | 0.95 |
| Recall (sensitivity) | 0.83 | 0.86 | 0.99 |
| F1-score | 0.86 | 0.86 | 0.97 |
| F2-score | 0.85 | 0.86 | 0.98 |

*Accuracy is derived by comparing to the gold standard set, the manually annotated notes. WCM refers to Weill Cornell Medicine, MSHS refers to Mount Sinai Health System, and UUHSC refers to*
University of Utah Healthcare Centre. All scores are a macro-average of the 0 and 1 categories. *F1* represents an equal balance between precision and recall while *F2* gives more weight to recall.

### Comparison of NLP and diagnostic codes

Overall, for the 14,462 patients in the WCM cohort, the validated NLP identified 3,245 (22.4%) patients with SI, whereas ICD codes identified 311 (2.2%), a greater than tenfold higher ascertainment rate compared with ICD codes (Table 3). The algorithm identified SB/SA at a rate 28-fold higher than that of ICD codes, with 74 (0.5% of the cohort) patients detected by ICD codes and 2,082 (14.4%) by NLP. When patients with suicide symptoms were identified by ICD codes, 99.1% of these patients were also identified by NLP (Table 4a), providing evidence that the algorithm discerned the same SI and SB/SA phenomena as the ICD codes, but for SB/SA the NLP outperformed diagnostic codes by detecting 28 times more cases because this information could mostly be detected in the free text notes.

**Table 3:** Overall ICD versus NLP Ascertainment.

|  | MSHS |  |  | WCM |  |  | Mean |
| --- | --- | --- | --- | --- | --- | --- | --- |
|  | ICD | NLP | NLP/ICD | ICD | NLP | NLP/ICD | NLP/ICD |
| **SU** | 2309 | 24108 | 10.44 | 339 | 3737 | 11.02 | 10.73 |
| **(%)** | (6.8%) | (71.3%) |  | (2.3%) | (25.8%) |  |  |
| **SI** | 1760 | 20482 | 11.64 | 311 | 3245 | 10.43 | 11.04 |
| **(%)** | (5.2%) | (60.6%) |  | (2.2%) | (22.4%) |  |  |
| **SB/SA** | 681 | 18440 | 27.08 | 74 | 2082 | 28.14 | 27.61 |
| **(%)** | (2.0%) | (54.6%) |  | (0.5%) | (14.4%) |  |  |

*SU* refers to Suicidality– either suicidal ideation (*SI*) or suicide-related behaviors or suicide attempt (*SA*). *NLP* refers to the Natural Language Processing algorithm described in the paper; *ICD* refers to the International Classification of Diseases diagnostic codes. *NLP/ICD* refers to the ratio of ascertainment of suicide term by *NLP* as compared to *ICD* at the sites: Mount Sinai Hospital System (*MSHS*) and Weill Cornell Medicine (*WCM*). The percentage refers to the number of patients identified out of 14,462 at *WCM* and 33,797 at *MSHS*.

**Table 4:**
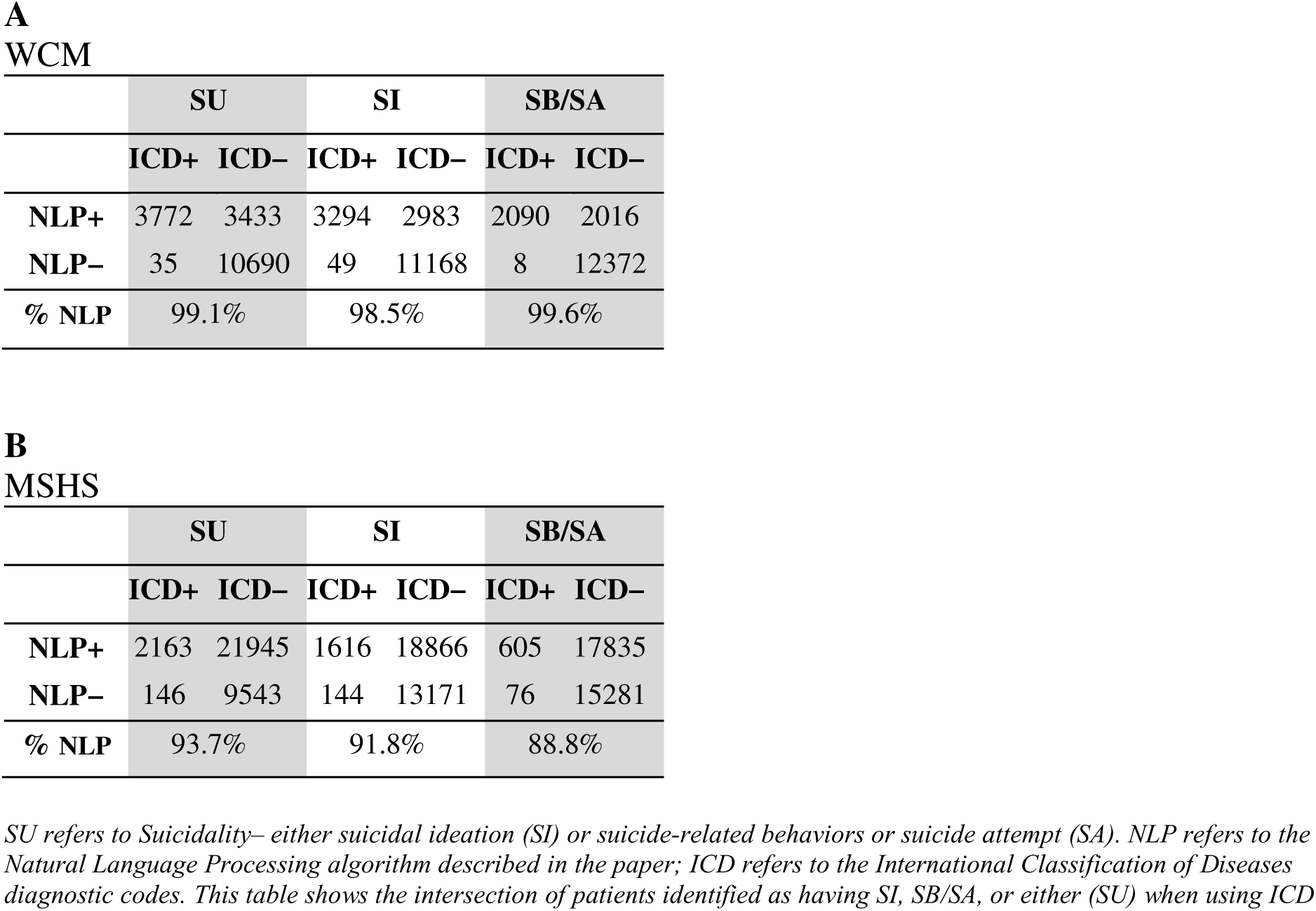
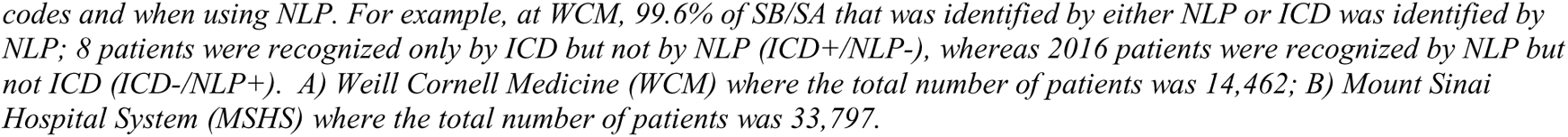
Overlap of ICD and NLP in ascertainment of suicidality.

Of the 33,797 patients in the MSHS cohort, 1,760 (5.2%) received ICD diagnoses for SI whereas 20,482 (60.6%) were detected by NLP, over an elevenfold higher ascertainment rate. ICD codes in MSHS records detected SB/SA in 681 (2.0%) patients, compared with NLP detecting SB/SA in notes of 18440 (54.6%) patients, a 27-fold higher ascertainment rate with NLP. Reassuringly, 94% of patients with a suicide related ICD code were also detected by NLP (Table 4b), suggesting that the algorithm not only identified patients with the relevant diagnostic codes, but also 27 times more patients with SB/SA because those findings were reported only in the free text notes.

At both sites, this pattern of NLP detection of SI being 10 fold higher and detection of SB/SA being 27-28 fold higher compared with ICD codes, was relatively comparable across different demographic groups (Supplemental Table S4 and S5). Notable exceptions included the race category mixed/other at WCM where the NLP rate for SI was 23 and SB/SA was 50 times that of the ICD code rate. As expected, because only a subset of individuals reporting SI also manifest SB/SA (18, 19), the overall prevalence of SB/SA (ICD or NLP) was lower than that of SI. Since SB/SA is the cause of more morbidity and predicts more mortality (2, 20) it is noteworthy that NLP identification is about three times better compared with diagnostic codes for SB/SA than it is for SI. Notable deviations involving even more advantage for NLP over diagnostic codes for SB/SA were observed in the 80+ year age category with a 72-fold superior NLP detection (MSHS) for, Black/African American with a 40-fold NLP advantage (MSHS), White with a 33- fold NLP advantage (MSHS), and widowed with a 71-fold NLP advantage (WCM). The disparity between NLP and ICD identification of SI and of SB/SA was smaller for a few psychiatric diagnoses including eating disorders, personality disorders, and posttraumatic stress disorder.

To investigate a bias effect on results due to the number of notes per case, we normalized the distribution of the number of notes per patient, and implemented a low-medium-high group stratification based on logarithmic means. Patients with more notes—and thus more opportunity for evaluation and documentation— were more likely to receive both ICD codes and free-text mentions of SI and SB/SA, but this did not explain our main findings of robustly higher ascertainment rates with NLP (Supplemental Tables S4 and S5).

### Performance across race and ethnicity groups

Due to the limited number of patients (Table 1), racial and ethnic demographic characteristics were grouped broadly into white, non-white, Hispanic/Latinx, and non-Hispanic/Latinx. The accuracy of the NLP pipeline maintained an F1 score of 0.80-0.87 for SI and 0.83-0.92 for SB/SA across groups at WCM and MSHS is reported (Supplemental Table S2a: WCM; S2b: MSHS).

### Performance outside psychiatric encounters

The note corpus at UUHSC, was divided to observe whether accuracy performance generalized beyond psychiatric notes to non psychiatric notes. Performance was comparable in a subset of 71 manually reviewed non-psychiatric notes with F1 scores 0.9 for detecting SI and 0.89 for detecting SB/SA in non-psychiatric notes (Supplemental Table S3).

## 4. Discussion

A portable NLP algorithm, based on the C-SSRS, was developed to detect SI and SB/SA from free text clinical notes in EHR of patients with psychiatric conditions. F1 scores for SI and SB/SA found in clinical notes were above 0.85 at each of the three test sites indicating validity of the method. This NLP detected 27-28 fold more SB/SA and 10 fold more SI compared with diagnostic codes in two medical centers. These scores are a significant improvement upon previous NLP applications in terms of portability across sites (16) and results were comparable to single-site studies (6, 7, 14).

Importantly, this NLP algorithm was developed based on the C-SSRS to accurately distinguish between ideation and behavior. Progress in preventing suicide has been limited by insufficient understanding about the transition from suicidal ideation to attempt (25). In what is known as the ideation-to-action framework, the development of SI is viewed as a process distinct from the progression of SI to SB/SA (26). Therefore, future EHR-based suicide research including prediction models, epidemiological studies, effectiveness research etc., can make the most impact by addressing ideation and behavior separately, as can be accomplished using this algorithm.

SB/SA is a stronger predictor of future suicidal behavior and suicide death (2, 3) and the fact that we found that the NLP algorithm had an ascertainment rate for SB/SA over diagnostic codes that was about three times better than its 10-fold advantage for SI over diagnostic codes, means that the need to use NLP instead of diagnostic codes is so much greater for the more serious form of suicidality, namely SB/SA.

Furthermore, the algorithm maintained performance in identifying both SB/SA and SI in minoritized racial and ethnic groups, which was not always observed in other reported NLP algorithms (21, 22). It was notable that the NLP advantage over diagnostic codes was even better in some race/ethnicity groups, suggesting a bias in allocation of diagnostic codes. The degrees of demographic bias in SI and SB/SA ICD codes is largely unknown, but its presence has previously been noted (23, 24). Our study provides some of the first evidence of such biases in diagnostic code allocation.

Results from UUHSC (Supplement Table S3) supported the utility of the algorithm outside psychiatric specialty notes.

An additional advantage of the lexicon approach over a machine-learning approach is the strict rule-based adherence to the C-SSRS. Language models, trained on very large text data, acquire their own inferred understanding of concepts such as suicidal symptoms which must be fine-tuned to perform the given task. Our previous work comparing the identification of mental health concepts using LLMs versus lexicon approaches demonstrated how LLMs may adhere to baseline definitions of concepts even after being fine-tuned (27). This may lead to a more colloquial classification of suicidal symptoms. Thus, a strict rule-based definition of suicidal outcomes based on the C-SSRS may be more clinically useful. On the other hand, the finite nature of the lexicon causes some mentions to be missed, or complex contexts to not be regarded. For example, in the manual error analysis, the algorithm missed “reports that [she] might be better off dead” as a mention of SI and “patient maintained on suicide precaution” which implies ongoing SI. Rare templates originally missed and thus not removed caused cases of false-positive (e.g. “call 911 or return to the nearest ER if unsafe, suicidal, homicidal”). Additional false positive examples included where the NegEx was unable to detect the negative context, in one case because “denies” SI was spelled incorrectly, and in another case due to the syntax: “does not say that he wants to die.” Some of these errors may be avoided by future work fine-tuning LLMs with C-SSRS classification examples. Our previous work comparing an LLM to a rules-based lexicon approach in a different nuanced psychiatric classification task illustrates the unique strengths and limitations of each approach (27). However, given the issues inherent to fully understanding deep learning model representations, our rule-based approach allows us to clearly identify the source of model errors, including clinical text misspellings or context; an LLM with millions of parameters may develop classification rules that are more difficult to understand, and thus, more difficult to debug. It is also of note that false negatives in ascertainment of SI and SB/SA have more severe clinical consequences than false positives. Future work regarding LLM training must be carefully balanced to maximize sensitivity of the model, as is accomplished in this algorithm.

The large-scale deployment study builds confidence for future users of this publicly available and user-friendly NLP SI and SB/SA detection approach. To help establish convergent validity and further detect bias, results of the NLP algorithm were compared to SI and SB/SA ICD code assignment. Given that more than 85% of cases with ICD-detected suicidality codes at both sites were also detected by NLP, provides confidence regarding the clinical construct captured by the NLP pipeline. Upon manual review, while there were some false-negatives, others occurred when the set of notes seen by the algorithm did not contain a clear positive mention of SI or SB/SA. It is possible that SI or SB/SA was mentioned in notes outside the corpus.

As expected, when divided into groups based on low, medium, and high numbers of notes, there were increasing ICD and NLP suicidality detection with more clinical notes. Detection by diagnostic codes also improved with more clinical notes, but the advantage of NLP over diagnostic codes did not change. This set of observations may mean that each encounter with a provider represents an additional opportunity for suicidality to be detected and recorded. It may also reflect that patients with greater suicidality are higher utilizers of the healthcare system (28), or that patients with more serious symptoms are more likely to be hospitalized, and thus, receive more intensive assessment and intervention.

When parsing by patients with certain psychiatric symptoms known to be associated with suicidality, the ICD prevalence was in the direction of more concordance with NLP. Certain groups stood out as being detected by NLP at even higher rates relative to diagnostic codes such as non-white race categories with up to double the SI ascertainment advantage for NLP, twice as high in the widowed marital status category, and almost three times as high in the 80+ age category (Supplemental Table 1a,b). These examples may reflect biased under-coding that is corrected by NLP, enabling these patients to be identified as having suicidal symptoms. Indeed, previous work in certain clinical populations has suggested a possible under-diagnosis of suicide ideation in patients of color (23), but also that the accuracy of billing codes and claims data is understudied (29). Finally, ICD codes do not necessarily distinguish between non-suicidal self harm and suicidal behaviors (30), adding to the problematic usage of ICD codes for EHR-based suicide research.

There were several limitations. To continue to improve the validity, fairness, and generalizability of the pipeline, bias detection (accuracy metrics as compared to gold-standard manual review) should be performed with a larger number of patients and stratified beyond white/non-white and Hispanic/non-Hispanic. Accuracy of the pipeline was lowest for detecting SI at MSHS in the white cohort (Appendix Table 4b). A limitation to understanding and improving the score was this group’s sample size of only ten patients. Future work that focuses on bias detection should utilize larger, balanced groups. Additional future directions may include consideration of more fine-grained phenotype categories such as ideation with or without plan and intent.

It should also be noted as a limitation when interpreting results of the ICD-NLP ascertainment comparison study that the medical records in this aim are not manually reviewed, so the proportion of false-positive NLP detection is unknown.

Language models leveraging sentence context might achieve higher predictive accuracy when compared to this rule-based approach (31). However, our results demonstrate that custom lexicons and task adaptation can produce modeling solutions with impressive classification capabilities without the need for intensive parameter fine-tuning. In contrast to many machine-learning based NLP approaches such as with Large Language Models (LLMs), this algorithm has ultimate transparency, explainability, and privacy– key tenants of responsible design and implementation of artificial intelligence technologies in healthcare (32). This is accomplished by using easy to interpret and understand lexicons of terms that can be viewed and tailored; no sensitive clinical note training data is required to input to a model. Finally, a lexicon-based approach is readily scalable across the range of healthcare systems rather than limited to use only within departments with computational infrastructure. The only modifications that may be required of future users are: 1) minor editing of lexicons (adding/removing words or phrases) to better suit the clinical notes in a particular setting; 2) collection and removal of clinical note templates to avoid irrelevant mentions of SI and SB/SA. Both are described in the Methods and can be accomplished without advanced data science skills or experience.

In summary, reliance on diagnostic codes for the detection of SI and SB/SB in EHRs results in substantial under-ascertainment of both outcomes. Future research with EHRs should not rely on diagnostic codes and employ an ascertainment strategy that differentiates SI and SA/SB. This shared resource for extracting SB/SA and SI from psychiatric clinical notes, was validated, is available for general use but can continue to be improved. The plan for the governance of GitHub is to continue iterative updating of the lexicons with feedback from implementation in a variety of note types at ours and other sites. Finally, the NLP detected a bias in diagnostic codes related to race/ethnicity that was not present in the NLP.

## Data Availability

The Electronic Healthcare Record data used in this study is not available given patient privacy and protections

## Acknowledgements

This study was supported by the National Institute of Mental Health (NIMH) grants: R01MH121921, R01MH121922, R01MH121923, and R01MH121924, and R01MH123489 and RF1MH134649.

We gratefully acknowledge the assistance of Dr. Samir Abdelrahman in the implementation of the NLP at the UUHSC site.

This work was supported in part through the computational and data resources and staff expertise provided by Scientific Computing and Data at the Icahn School of Medicine at Mount Sinai and supported by the Clinical and Translational Science Awards (CTSA) grant UL1TR004419 from the National Center for Advancing Translational Sciences. Research reported in this publication was also supported by the Office of Research Infrastructure of the National Institutes of Health under award number S10OD026880 and S10OD030463. The content is solely the responsibility of the authors and does not necessarily represent the official views of the National Institutes of Health.

At the UUHCS site, data linking was made possible through the Utah Population Database (UPDB). Partial support for all datasets housed within the UPDB is provided by the Huntsman Cancer Institute (HCI), http://www.huntsmancancer.org/, and the HCI Cancer Center Support grant, P30CA42014 from the National Cancer Institute. Research was supported by NCRR grant “Sharing statewide health data for genetic research” R01RR021746 with additional support from the Utah Department of Health and Human Services and the University of Utah. We thank University of Utah Health Data Science Services for data and analytics support, and the University of Utah Pedigree and Population Resource and the University of Utah Health Enterprise Data Warehouse for establishing the Master Subject Index between the Utah Population Database and the University of Utah Health Sciences Center. We also thank Drs. Brooks Keeshin, Brent Kious, Anna Docherty, Hunter Strohmeyer, and Michael Pope for their assistance in manual validation of the NLP algorithm, and Dr. Samir Abdelrahman for his technical assistance in implementing the algorithm.

