## Supplemental tables for "A Natural Language Processing Pipeline based on the Columbia-Suicide Severity Rating Scale"

**Supplemental Table S1. Cohen’s Kappa Interrater Agreement**

|  | WCM (n=400) | MSHS (n=327) | UUHSC (n=404) |
| --- | --- | --- | --- |
| Suicidal ideation | 0.79 | 0.82 | 0.75 |
| Suicidal behavior | 0.78 | 0.93 | 0.72 |

**Supplemental Table S2: Accuracy metrics of the NLP algorithm across race/ethnicity groups**

**A Weill Cornell Medicine**

| **Demographic Category (number of patients)**  **Number of notes** | **Non-white (n=11)**  129 notes | **White (n=21)**  271 notes | **Hispanic/Latinx (n=2)**  30 notes |
| --- | --- | --- | --- |
| **Suicidal ideation** |  |  |  |
| Accuracy | 0.91 | 0.94 | 0.90 |
| Precision (positive predictive value) | 0.86 | 0.85 | 0.78 |
| Recall (sensitivity) | 0.93 | 0.92 | 0.84 |
| F1-score  F2-score | 0.88  0.92 | 0.88  0.91 | 0.80  0.83 |
| **Suicidal behavior/attempt** |  |  |  |
| Accuracy  Precision (positive predictive value)  Recall (sensitivity)  F1-score  F2-score | 0.93  0.92  0.90  0.91  0.90 | 0.87  0.92  0.80  0.83  0.82 | 0.97  0.98  0.83  0.89  0.86 |

**B Mount Sinai Health System**

| **Demographic Category (number of patients)**  **Number of notes** | **Non-white (n=36)**  256 notes | **White (n=10)**  70 notes | **Hispanic/Latinx (n=9)**  67 notes |
| --- | --- | --- | --- |
| **Suicidal ideation** |  |  |  |
| Accuracy | 0.88 | 0.87 | 0.87 |
| Precision (positive predictive value) | 0.85 | 0.84 | 0.86 |
| Recall (sensitivity) | 0.9 | 0.87 | 0.87 |
| F1-score  F2-score | 0.87  0.89 | 0.85  0.86 | 0.86  0.87 |
| **Suicidal behavior/attempt** |  |  |  |
| Accuracy  Precision (positive predictive value)  Recall (sensitivity)  F1-score  F2-score | 0.85  0.85  0.85  0.85  0.85 | 0.87  0.88  0.87  0.87  0.87 | 0.93  0.91  0.93  0.92  0.93 |

*S2A refers to WCM data while S2B refers to MSHS data. Metrics are presented along the rows stratified by outcome, and the number of notes are denoted in the column headers. The numbers at the top of the columns correspond to the total number of notes per demographic. All scores are a macro-average of the 0 and 1 categories.*

**Supplemental Table S3: Performance Metrics Stratified by Note Category at University of Utah Healthcare Centre (UUHSC)**

| **Category (number of notes)** | **Psychiatric (n=275)** | **Non-Psychiatric (n=129)** | **Total (n=404)** |
| --- | --- | --- | --- |
| **Suicidal ideation** |  |  |  |
| Accuracy | 0.99 | 0.91 | 0.96 |
| Precision (positive predictive value) | 0.99 | 0.83 | 0.95 |
| Recall (sensitivity)  F1-score  F2-score | 0.96  0.98  0.97 | 0.97  0.90  0.94 | 0.97  0.96  0.97 |
| **Suicidal behavior/attempt** |  |  |  |
| Accuracy | 0.97 | 0.95 | 0.96 |
| Precision (positive predictive value) | 0.93 | 0.92 | 0.91 |
| Recall (sensitivity)  F1-score  F2-score | 0.95  0.94  0.95 | 0.87  0.89  0.88 | 0.99  0.95  0.97 |

*SI here refers to suicidal ideation, and SBSA refers to suicidal behaviour/suicidal attempt. The Psychiatric column denotes clinical notes from psychiatric encounters while Non-Psychiatric are notes from non-psychiatric encounters. All scores are a macro-average of the 0 and 1 categories.*

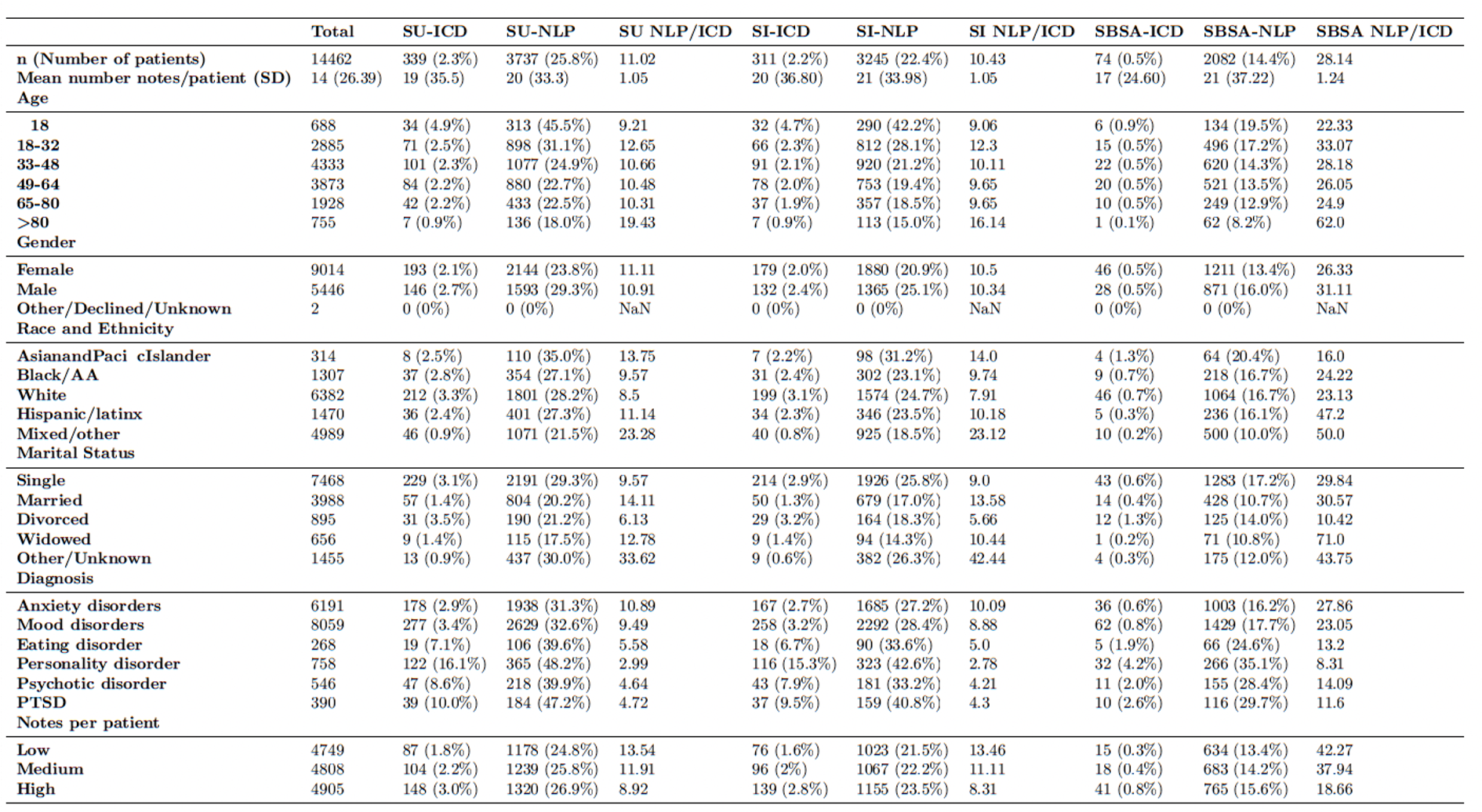
**Supplemental Table S4: ICD and NLP Ascertainment at WCM**

*ICD columns refer to the numberof patients obtained using ICD codes, NLP columns refer to the number of patients obtained using NLP algorithms. Low, medium, high refer to the number of patients with a low (1-6),medium (7-12), and high (13-200) number of psychiatric notes in their EHR.*

**Supplemental Table S5: ICD and NLP Ascertainment at MSHS**

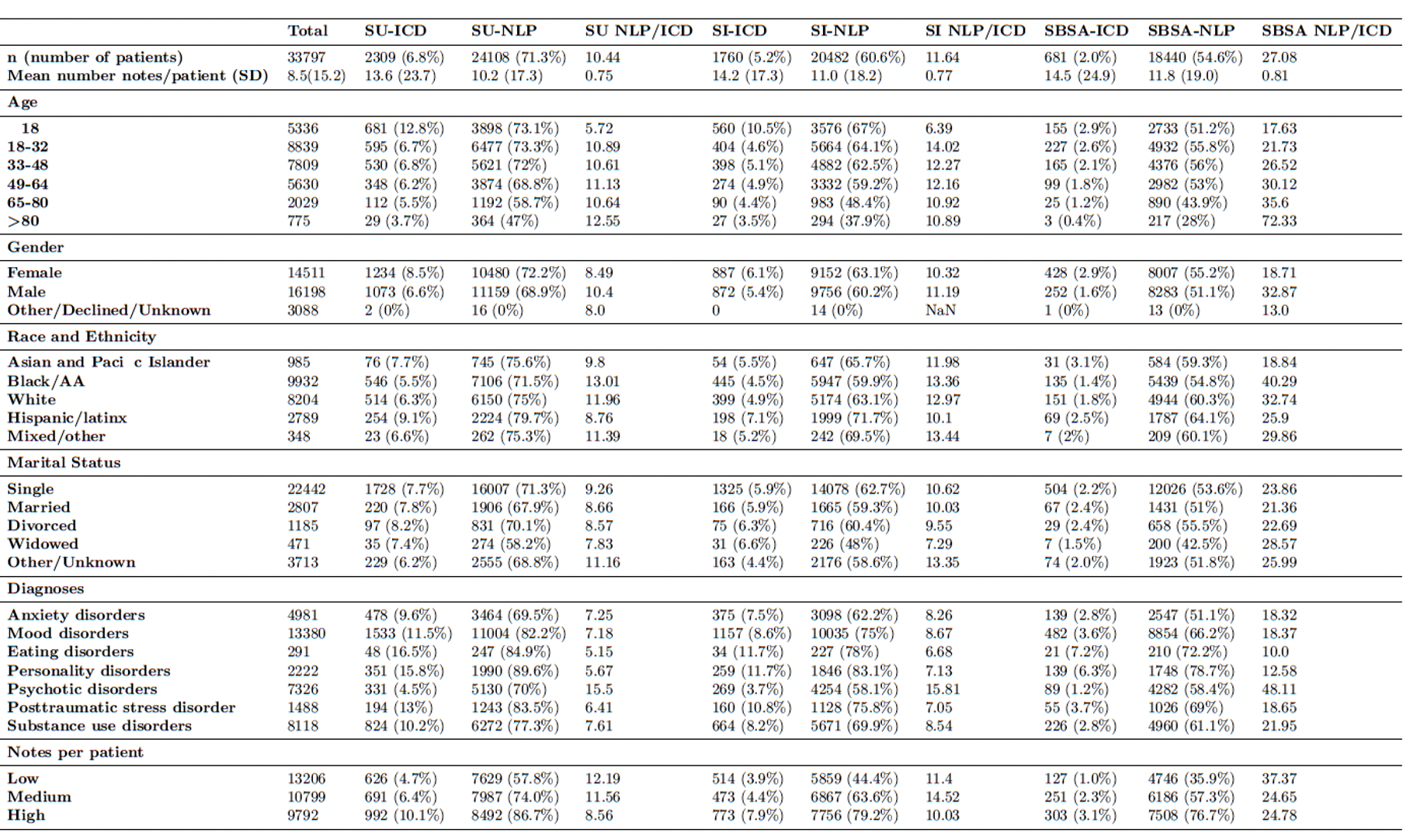

*ICD columns refer to the number of patients obtained using ICD codes, NLP columns refer to the number of patients obtained using NLP algorithms. Low, medium, high refer to the number of patients with a low (1-6),medium (7-12), and high (13-200) number of psychiatric notes in their EHR.*
