## Supplement 1 for "A Natural Language Processing Pipeline based on the Columbia-Suicide Severity Rating Scale"

WCM is an academic medical center in New York City. Data were derived from patients with clinical notes authored by clinicians from multiple clinical specialties between 2000 and 2020. To increase the likelihood of assessing patients with psychiatric symptoms, the structured EHR data were used to narrow the cohort to 177,993 patients who were prescribed one or more psychotropic medication and further to 14,462 patients with at least one encounter with a psychiatric provider. This study was conducted under WCM IRB# 1510016639.

The MSHS in New York City provides care to a diverse patient population. To select a distinct population from that of WCM, the corpus was narrowed to visits from specific high-acuity care settings in which suicidality was very likely to be evaluated: inpatient psychiatry, psychiatric emergency departments, and inpatient psychiatric consult-liaison encounters between 2011 and 2020 at three hospital locations. This corpus consisted of 286,459 notes from 33,800 patients in the MSHS Data Warehouse. This work was performed under the approved IRB Protocol Study-20-00338 at the Icahn school of Medicine at Mount Sinai.

The UUHSC is the only academic medical center in the Mountain West region, providing patient care for Utah, Idaho, Wyoming, Montana, western Colorado, and Nevada. For this cohort, the goal was to assess the accuracy of the pipeline in patients who died by suicide and had previous ICD codes indicating a history of prior nonfatal SI/SB (n=22 patients and 6,249 notes) compared to living individuals with at least two ICD codes for affective disorders but no ICD codes for suicidal thoughts or behaviors(n=22 patients and 9,099 notes). This specific cohort was selected to investigate whether the NLP pipeline would detect suicidality where it was expected (suicide deaths with ICD codes indicating prior nonfatal SI/SB) vs. where it was not apparent in the billing codes (living individuals with no codes for SI/SB). The two groups were matched on sex, age, and race. Clinical notes were evaluated across specialties. This work was performed under the approved IRB Protocol Study, Utah IRB_00133374.
