## Supplement 2 for "A Natural Language Processing Pipeline based on the Columbia-Suicide Severity Rating Scale"

At MSHS, templates such as "Is the patient at elevated risk for suicidal behavior in the next 24 hours?" can be found in clinical notes. While this template does not answer whether the patient actually presented with suicidal behavior, the presence of the phrase “suicidal behavior” would result in a false-positive detection. Templates were iteratively collected and removed from the note, and only free text mentions of suicidal symptoms were considered.

At UUHSC, where the notes were not all written by psychiatric providers, the common abbreviations “SI” and “SA” were removed from the lexicons to minimize false-positive results from notes where these phrases were used to characterize medical outcomes.
